# Uncovering the Prevalence of Cystinosis through Genetic Analysis

**DOI:** 10.1101/2023.05.04.23289520

**Authors:** Chen-Han Wilfred Wu, Alicja Tomaszewski, Louisa A. Stark, Fernando Scaglia, Ewa Elenberg, Fredrick R. Schumacher

## Abstract

**Background:** Cystinosis is a metabolic disease characterized by the accumulation of cystine most often presenting in an infantile nephropathic form caused by pathogenic variants in the *CTNS* gene. It is characterized by progressive loss of glomerular function leading to renal failure by the first decade of life, making early diagnosis crucial to improving outcomes. This study seeks to estimate the prevalence of cystinosis using a population genetics approach.

**Methods:** The Human Genome Mutation Database (HGMD) was used to identify known pathogenic variants in *CTNS*, and the 1000 Genomes (1KG) database was used to identify *CTNS* variants in a cohort representing a healthy population. These two databases were intersected to identify disease-causing variants and their carriers in the general population. The Hardy-Weinberg equilibrium was used to calculate expected carrier and affected rates for cystinosis.

**Results:** The allele frequency for all disease-causing alleles was calculated to be 0.016. The predicted affected rate was calculated to be 0.00027 (approximately 1:3680), and the predicted carrier rate was 0.032 (approximately 1:30).

**Conclusion:** Compared to the reported clinical prevalence of between 1 in 100,000 to 1 in 200,000, the prevalence of cystinosis in this study was calculated to be 1 in 3,680. This significantly higher result may be due to the underdiagnosis of cystinosis or variable expressivity of variants presenting with a broad range of disease severity. These results support the proposal of newborn screening for early diagnosis and improved outcomes in infantile nephropathic cystinosis.

## Introduction

Cystinosis is a rare autosomal recessive disorder that is characterized by systemic accumulation of cystine crystals in different types of cells and tissues. It is also the most common inherited cause of Fanconi syndrome observed in children.[1] Cystine crystals begin to accumulate at low pH due to impaired transport out of lysosomes, and symptoms begin to appear several months after birth.[2, 3] Some of the associated symptoms include polyuria as well as urinary loss of low-molecular weight protein, glucose, amino acids, phosphate, calcium, magnesium, sodium, potassium, bicarbonate, carnitine, and water.[2] In late childhood, many patients with cystinosis can develop medullary nephrocalcinosis.[4] Nephropathic cystinosis has been identified as a metabolic disease causing Fanconi syndrome which predisposes affected individuals to nephrolithiasis, especially among the pediatric population.[5] Although symptoms of cystinosis are often associated with renal function, cystine crystal accumulation occurs in most cells and tissues of the body including the conjunctiva, corneas, liver, spleen, lymph nodes, kidneys, thyroid, intestines, rectal mucosa, muscle, brain, macrophages, and bone marrow.[2] Without timely treatment, children with cystinosis will go on to develop photophobia due to cystine crystal accumulation in the cornea as well as hypothyroidism due to the accumulation of cystine in thyroid tissue at around 10 years of age.[6] This makes early diagnosis critical to limit the damaging manifestations of cystinosis.

The epidemiology of cystinosis varies globally, ranging from 1 in 115,000 in Denmark to 1 in 260,000 in Sweden. Several populations have been identified that suggest an increased incidence rate due to founder mutations or in communities in which consanguinity is more common.[7] There are an estimated 500-600 patients in the United States diagnosed with cystinosis, corresponding to a prevalence of 1 in 479,000 to 1 in 575,000.[2, 8]

Cystinosis is a monogenic disease that is caused by pathogenic variants in the *CTNS* gene located on chromosome 17p13.[9] *CTNS* encodes the carrier membrane protein cystinosin, which aids in the transport of cystine out of the lysosomal compartment. Deficiency or defect in cystinosin causes cystine to build up in the lysosomes of various organ tissues in the body. This accumulation of cystine results in crystal formation, especially in the proximal tubules of nephrons, which manifests as the nephropathic presentation of cystinosis.

Cystinosis is classified into three distinct clinical presentations: the infantile nephropathic form, the juvenile nephropathic form, and the ocular non-nephropathic form. The most common and severe presentation is the infantile nephropathic form, accounting for 95% of cases of cystinosis. This form most often leads to progressive loss of glomerular function ultimately leading to complete renal failure by the end of the first decade of life.[10, 11] A smaller proportion of patients (5%) are diagnosed later in childhood with the juvenile form of cystinosis and show milder pathologies compared to the infantile nephropathic form.[12] The non-nephropathic ocular form is rare and characterized by corneal cystine accumulation resulting in photophobia, and it presents in adulthood.[13]

Currently, diagnosis of cystinosis relies primarily on measuring leukocyte cystine content or the presence of corneal crystals on slit-lamp examination; however, crystals may not form until one year after birth.[14] Cystinosis can also be confirmed by genetic assays identifying variants in the *CTNS* gene. Studies have shown that diagnosing cystinosis at an early age by integrating it into newborn screening programs followed by appropriate treatment is critical to limit the systemic damage caused by the disease.[15]

This study seeks to better understand the genetic prevalence of cystinosis by using population databases of genetic information. Additionally, this study will help identify specific variants that are carried by the general population to earlier diagnosis and treatment of this disease. By facilitating earlier diagnosis, the renal and systemic damage associated with the disease can be limited or prevented in the young population that it predominantly impacts.

## Methods

Pathogenic variants in CTNS that are known to cause cystinosis were collected from the Human Gene Mutation Database (HGMD) v.2012.3. Types of variants in this gene included missense, nonsense, insertions, and deletions. The 1000 Genomes (1KG) database was used to identify CTNS variants in a cohort representing a healthy population (Table 1). This database consists of genomes collected from a representative global population with 98% of alleles with a frequency of more than 1% in the populations that it samples from, in addition to alleles that have a lower frequency.[16] The list of pathogenic variants from the HGMD was intersected with the variants carried in the 1KG database to identify disease-causing variants and their carriers in the general population (Figure 1). All related individuals were excluded from the analysis.

**Table 1.**
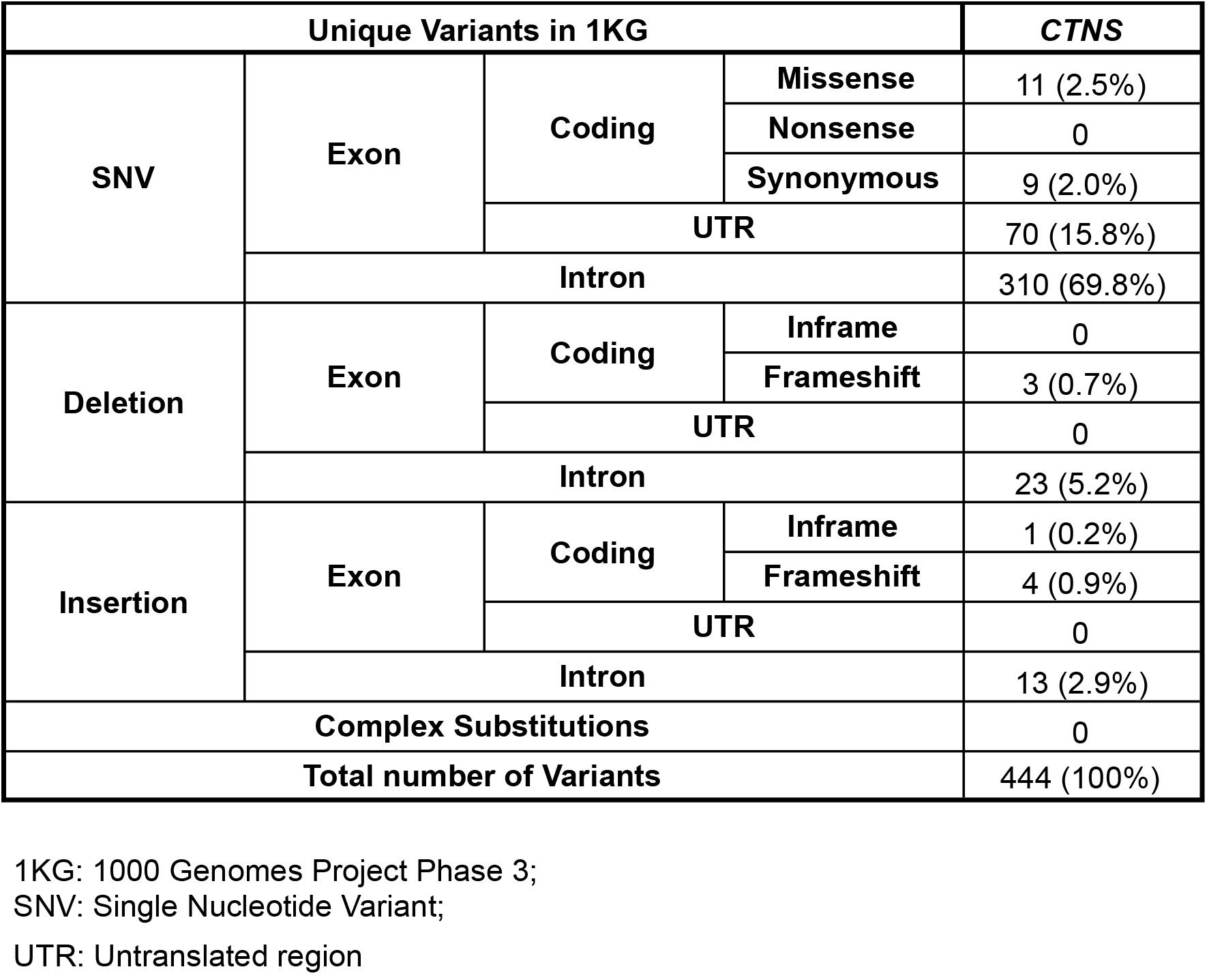
Classification of CTNS unique variants in the 1KG database.

**Figure 1:**
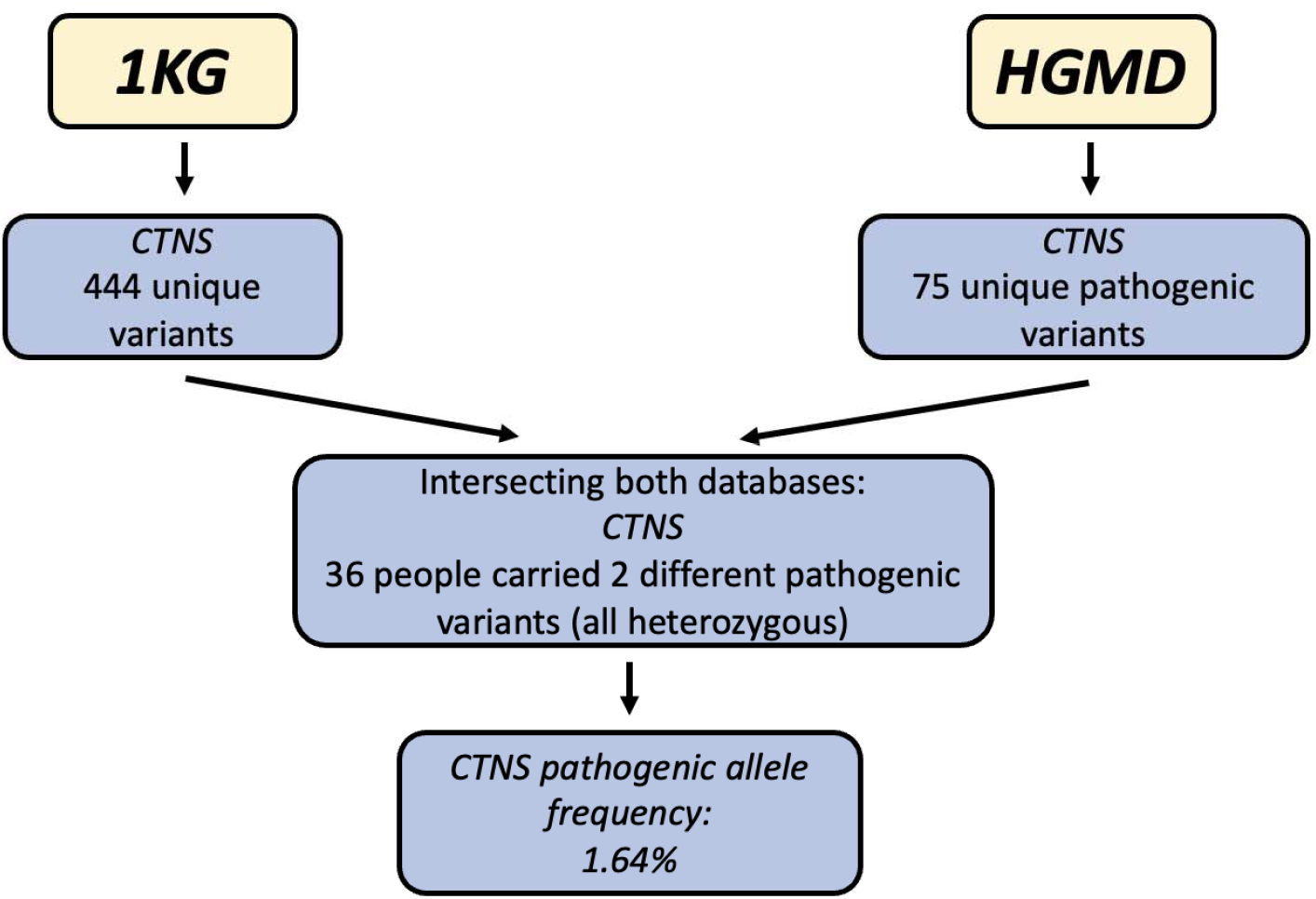
Flowchart showing population genetics method applied in study using Human Genome Mutation Database (HGMD) to identify known pathogenic *CTNS* variants and 1000 Genomes (1KG) database to identify individuals with *CTNS* variants in a healthy representative population. There were 75 unique pathogenic variants in HGMD and 444 unique variants in 1KG. After intersecting the two databases, there were 36 people sharing 2 different pathogenic variants.

Using the 1KG database, pathogenic allele frequencies (q) were estimated based on individuals harboring the pathogenic variant. Expected carrier and affected rates were calculated using the Hardy-Weinberg equilibrium (p^2^ + 2pq + q^2^ = 1) (Figure 1).

## Results

*CTNS* variants were identified using the HGMD and 1KG databases. Using the 1KG database, 444 unique *CTNS* variants were found among 1092 healthy individuals, as demonstrated in Supplemental Table 1. The majority of these were single nucleotide variants (SNV) in the intron regions (69.8%). Other variants including missense, synonymous, and SNVs in the UTR, as well as deletions and insertions, are shown in Table 1. Most insertions and deletions were observed in the intron regions (5.2% deletions, 2.9% insertions) compared to exon regions (0 deletions, 0.7% insertions). The lack of nonsense or stop-gain variants in the 1KG in this gene suggests that these types of mutations are deleterious to the function of the gene and cannot be sustained in a healthy individual.

The HGMD database was used to identify variants that are known to be associated with a pathology or disease. Using HGMD, 75 pathogenic *CTNS* variants were identified including missense, nonsense, insertions, deletions, and complex substitutions. The variants identified were mostly SNV missense variants (45.3%) as well as exon deletions (9.3% in-frame, 21.3% frameshift), as shown in Table 2. This contrasts with the variants that were observed in the 1KG database, which were primarily intron SNV variants. Three complex substitutions were identified in the *CTNS* gene that show multiple consecutive nucleotides with significant alterations (Table 2).

**Table 2.** Classification of CTNS unique variants in the HGMD database.

| Unique Pathogenic Variants in HGMD |  |  |  | CTNS |
| --- | --- | --- | --- | --- |
| SNV | Exon | Coding | Missense | 34 (45.3%) |
|  |  |  | Nonsense | 7 (9.3%) |
|  |  |  | Synonymous | 0 |
|  |  | UTR |  | 0 |
|  | Intron |  |  | 0 |
| Deletion | Exon | Coding | Inframe | 7 (9.3%) |
|  |  |  | Frameshift | 16 (21.3%) |
|  |  | UTR |  | 0 |
|  | Intron |  |  | 0 |
| Insertion | Exon | Coding | Inframe | 1 (1.3%) |
|  |  |  | Frameshift | 7 (9.3%) |
|  |  | UTR |  | 0 |
|  | Intron |  |  | 0 |
| Complex Substitutions |  |  |  | 3 (4%) |
| Total number of Pathogenic Variants |  |  |  | 75 (100%) |

HGMD: Human Gene Mutation Database;
SNV: Single Nucleotide Variant;
UTR: Untranslated Region

The HGMD and 1KG databases were intersected and identified 2 disease-causing variants that were present in both datasets (Table 3). A total of 36 individuals carried these 2 disease-causing variants (Table 3). A total of 35 individuals were shown to carry the c.124G>A variant identified in both databases, and the c.473T>C variant identified in both datasets was carried by 1 individual in the 1KG database. All the individuals with these variants were heterozygous carriers, with no homozygotes, compound heterozygotes, multiple pathogenic variants in cis or trans, double homozygotes, or double heterozygotes. Specific characteristics of these two variants including the chromosomal location, position and exon number, reference and variant alleles, and observed allele frequency are found in Table 3.

**Table 3.** Characteristics of intersected genetic variants (pathogenic variants) of CTNS in 1KG and HGMD.

| Chromosome | Position | Variant Identifier | Reference Allele | Variant Allele | C. nomenclature | P. nomenclature | Exon number | Type | Number of people carrying this variant (all heterogenous) | Observed Allele Frequency (%) |
| --- | --- | --- | --- | --- | --- | --- | --- | --- | --- | --- |
| chr17 | 3550800 | rs35086888 | G | A | c.124G>A | p.Val42Ile | 4/13 | missense | 35 | 35/2184 (1.60%) |
| chr17 | 3559792 | rs113994206 | T | C | c.473T>C | p.Leu158Pro | 8/13 | missense | 1 | 1/2184 (0.04%) |

Chromosome position based on Genome Reference Consortium Human Build 37 (GRCh37, hg19)
1KG: 1000 Genomes Project Phase 3; HGMD: Human Gene Mutation Database SNV: Single Nucleotide Variant

Based on these results, the disease-causing alleles in the *CTNS* gene were found to have a frequency of 1.64% (Table 3, Table 4). The Hardy Weinberg Equilibrium (p^2^ + 2pq + q^2^ = 1), was used to determine expected carrier and affected rates for *CTNS* pathogenic variants. In this equation, q represents the frequency of disease-causing variants in *CTNS*, which was calculated to be 1.64%. Using this value, the expected carrier rate was calculated to be 1 in 30, and the affected rate was calculated to be 1 in 3,680. These findings are summarized in Table 4.

**Table 4.**
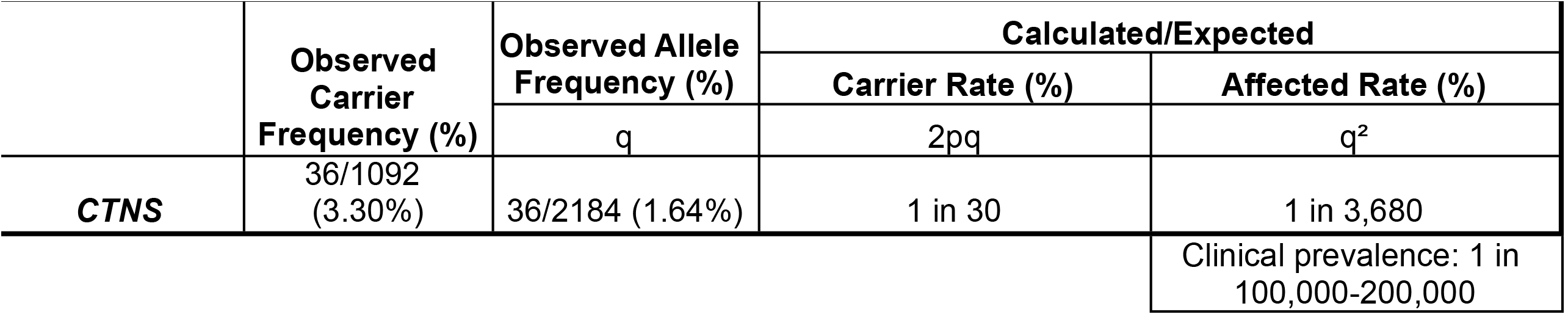
Observed and Calculated allele frequencies using Hardy-Weinberg equilibrium.

## Discussion

This study was designed to estimate the prevalence of cystinosis using the genetic approach of investigating pathogenic *CTNS* variants in the HGMD and 1KG databases. Of the 75 unique pathogenic variants observed in the HGMD database (Table 2), 2 unique variants were present in the 1KG database (Table 3). Both unique pathogenic variants observed in the intersection of these two databases corresponded to missense variants in the *CTNS* exon regions.

### Comparison between genetic prevalence and clinical prevalence

There are an estimated 500-600 patients in the United States diagnosed with cystinosis, corresponding to a prevalence of 1 in 479,000 to 1 in 575,000.[2, 8]

By contrast, the genetic approach used in our study estimated the overall rate of cystinosis to be 1 in 3,680 (Table 4), approximately 24x higher than the reported clinical incidence. There are various potential explanations for this discrepancy, including an underdiagnosis of cystinosis as well as variable expressivity of disease-causing variants.

### Underdiagnosis and support for genetic diagnosis by newborn screening

Based on the significant difference between the estimated prevalence of cystinosis in this study and the clinically reported value, there is a high likelihood that cystinosis is underdiagnosed. The specific clinical guidelines of cystinosis diagnosis include the identification of cystine crystals on slit lamp examination, elevated cystine levels in polymorphonuclear leukocytes, and increased cystine content in cultured fibroblasts or in the placenta at birth.[10] These approaches may have missed many potentially treatable cases of cystinosis. Our study supports the potential need of revising the diagnostic criteria to improve sensitivity.

While the presence of biallelic pathogenic variants in the *CTNS* gene is also one of the diagnostic criteria for cystinosis, it is not routinely performed. Early diagnosis of cystinosis is critical as this helps limit the irreversible damage that may be caused by cystine deposition, such as renal manifestations. Treatment options such as cystine-depleting agents exist for cystinosis, and outcomes are better for cases identified as soon as possible after birth. As discussed earlier, the estimated prevalence of cystinosis shown in this study is significantly higher than expected based on the reported clinical prevalence, suggesting that current clinical diagnosis guidelines may not be sufficient to identify all cases. These factors all contribute towards support for implementing genetic diagnosis, such as through newborn screening for cystinosis with genetic analysis, to capture more individuals with this diagnosis before they begin to show deleterious primary manifestations of the disease.

### Potential for variable expressivity

Another potential explanation for the results observed in this study is the possibility of variable expressivity of variants in the *CTNS* gene that cause cystinosis. After cross-referencing the 1KG and HGMD databases, only two *CTNS* variants from those listed in the HGMD were observed in the representative 1KG population. This suggests that most pathogenic variants found in HGMD do not exist in the general population. Some of these pathogenic variants may be incompatible with life and therefore may not be represented at a sufficiently high level to be observable at the population level; however, some pathogenic variants may have a milder presentation, and the frequency of these variants may be comparatively greater in the population. These propose the potential for variable expressivity of variants in *CTNS* that cause cystinosis. Further functional studies or longitudinal follow up clinically are needed to better delineate the severity of the disease that different variants cause.

### Strengths and Limitations

The major strength of this study is the use of HGMD as a source enumerating the known disease-causing variants in *CTNS* associated with cystinosis. This database is manually curated by experts in the field, allowing for a reliable estimate of the pathogenicity of the variants in the *CTNS* gene. The use of the 1KG database further strengthens this study as a source of individual level genetic data as opposed to aggregate data to determine carrier and allele frequencies. Using this individual level genetic data, we are able to confirm that there are no homozygotes, compound heterozygotes, or double heterozygotes.

Although the use of the HGMD and 1KG databases serves as a significant strength of the study, there are several weaknesses. The professional version of HGMD used in this study is proprietary and therefore not available for use by the public. Additionally, the 2014 version of HGMD was used in this analysis which may not include new pathogenic variants that were not captured in our genetic analysis. Since our current results already show a significantly increased estimated prevalence compared to what is clinically reported, new variants that were not included in our genetic analysis would further increase the genetic prevalence.

Finally, there may be a discrepancy in the representation of ethnic groups in genetic studies. Studies suggest that there is greater hesitancy among some ethnic minorities to utilize genetic analyses, which decreases the availability of clinically useful data for these groups.[17] The lack of a representative population can affect what is considered pathogenic, as some variants may be overrepresented in some groups over others. While HGMD and 1KG capture representative global populations, further studies are needed to assess potential differences across ethnic groups.

## Conclusion

This study aims to better characterize the prevalence of cystinosis by assessing disease-causing variants in the *CTNS* gene, which encodes cystinosin, the protein responsible for cystine transport out of the lysosomal compartment. Among the 75 pathogenic variants in *CTNS* identified using the HGMD database, only 2 of these variants were carried in the general population in the 1KG database. The genetic prevalence of cystinosis estimated by the study (1 in 3,680) was significantly higher than the reported clinical prevalence (between 1 in 479,000 to 1 in 550,000). This may be a result of underdiagnosis of cystinosis or variable expressivity of variants that present with a broad range of disease severity. The significant difference between our results and the clinically reported prevalence, combined with the global variability in reported cystinosis prevalence between countries, suggests that there is potential for improvement in the diagnosis of the disease. Based on our results, we would advocate for early genetic diagnosis, as early as newborn screening. The importance of diagnostic sensitivity is underscored by the improved outcomes with early treatment in cases of infantile nephropathic cystinosis. Further analysis is needed to improve our screening for this disease in addition to identifying additional disease-causing variants in the population.

## Data Availability

All data produced in the present study are available upon reasonable request to the authors

https://www.internationalgenome.org/

## Acknowledgements

We would like to express our deep gratitude to the late Dr. Lee-Jun Wong, PhD, a renowned biochemical and molecular geneticist at Baylor College of Medicine, for her invaluable contributions, insightful comments, and expert guidance in the design of our analysis. Her dedication and support were instrumental in the preparation of this study and manuscript. We are honored to have had the opportunity to collaborate with her. This manuscript is an expansion on a previously presented abstract at the American Society of Human Genetics 65th Annual Meeting in Baltimore, MD in October 2015.

## Ethics approval statement

### Research involving Human Participants and/or Animals

This is an observational study. The data used is de-identified and publicly available. The Case Western Reserve University/ University Hospital institutional review board has confirmed that no ethical approval is required.

### Informed consent

Given the deidentified nature of the data, the Case Western Reserve University/ University Hospital institutional review board determined that this observational study did not constitute human participants research and was thus exempted from review and the need for informed consent, in accordance with 45 CFR §46.

