## supplementary table 1 for "Uncovering the Prevalence of Cystinosis through Genetic Analysis"

**Supplementary Table 1. Variants of *CTNS* in the 1KG database**

| Chromosome | Start Position | End Position | Variant Type | Reference Genotype \| Proteotype | Variant 1 Genotype \| Proteotype | Count | Variant 2 Genotype \| Proteotype | Count |
|---|---|---|---|---|---|---|---|---|
| chr17 | 3539835 | 3539835 | SNV | T | C:C | 4 | C:T | 106 |
| chr17 | 3539897 | 3539897 | SNV | C | C:T | 102 | T:T | 4 |
| chr17 | 3540042 | 3540042 | SNV | G | A:A | 43 | A:G | 281 |
| chr17 | 3540136 | 3540136 | SNV | A | A:G | 47 | G:G | 2 |
| chr17 | 3540163 | 3540163 | SNV | A | A:G | 49 | G:G | 2 |
| chr17 | 3540662 | 3540662 | deletion | T | -:- | 1092 | | |
| chr17 | 3540693 | 3540693 | SNV | G | A:A | 169 | A:G | 439 |
| chr17 | 3540885 | 3540885 | SNV | C | C:T | 1 | | |
| chr17 | 3540917 | 3540917 | SNV | T | C:C | 171 | C:T | 441 |
| chr17 | 3541106 | 3541107 | deletion | TC | -:- | 3 | -:TC | 108 |
| chr17 | 3541107 | 3541109 | deletion | CTC | -:- | 4 | -:CTC | 102 |
| chr17 | 3541111 | 3541113 | deletion | TCA | -:- | 3 | -:TCA | 102 |
| chr17 | 3541113 | 3541113 | SNV | A | A:G | 23 | G:G | 3 |
| chr17 | 3541140 | 3541140 | SNV | C | C:T | 2 | | |
| chr17 | 3541143 | 3541145 | deletion | CTT | -:- | 8 | -:CTT | 94 |
| chr17 | 3541172 | 3541172 | deletion | A | -:- | 50 | -:A | 172 |
| chr17 | 3541208 | 3541208 | SNV | C | A:A | 4 | A:C | 102 |
| chr17 | 3541283 | 3541283 | SNV | C | A:C | 10 | | |
| chr17 | 3541350 | 3541350 | SNV | G | A:G | 30 | | |
| chr17 | 3541431 | 3541431 | SNV | G | G:T | 26 | | |
| chr17 | 3541441 | 3541441 | insertion | - | -:A | 144 | A:A | 16 |
| chr17 | 3541504 | 3541504 | SNV | T | G:T | 6 | | |
| chr17 | 3541766 | 3541766 | SNV | A | A:G | 283 | G:G | 41 |
| chr17 | 3541818 | 3541818 | SNV | C | C:T | 10 | | |
| chr17 | 3541940 | 3541940 | SNV | C | C:T | 23 | T:T | 3 |
| chr17 | 3542067 | 3542067 | SNV | G | G:T | 2 | | |
| chr17 | 3542075 | 3542075 | SNV | G | A:A | 170 | A:G | 442 |
| chr17 | 3542119 | 3542119 | SNV | A | A:G | 3 | | |
| chr17 | 3542136 | 3542136 | SNV | A | A:C | 315 | C:C | 43 |
| chr17 | 3542162 | 3542162 | SNV | C | C:T | 24 | T:T | 1 |
| chr17 | 3542166 | 3542166 | SNV | C | C:T | 17 | | |
| chr17 | 3542208 | 3542208 | SNV | G | A:G | 5 | | |
| chr17 | 3542243 | 3542243 | SNV | G | A:G | 3 | | |
| chr17 | 3542283 | 3542283 | SNV | G | C:C | 48 | C:G | 131 |
| chr17 | 3542455 | 3542455 | SNV | T | C:C | 170 | C:T | 441 |
| chr17 | 3542479 | 3542479 | SNV | G | A:A | 170 | A:G | 440 |
| chr17 | 3542485 | 3542485 | SNV | C | C:T | 207 | T:T | 65 |
| chr17 | 3542524 | 3542524 | SNV | C | C:G | 442 | G:G | 170 |
| chr17 | 3542593 | 3542593 | SNV | C | C:G | 442 | G:G | 170 |
| chr17 | 3542626 | 3542626 | insertion | - | -:T | 69 | T:T | 2 |
| chr17 | 3542709 | 3542709 | SNV | C | A:A | 2 | A:C | 35 |
| chr17 | 3542768 | 3542768 | SNV | G | C:C | 166 | C:G | 437 |
| chr17 | 3542819 | 3542819 | SNV | G | A:G | 6 | | |
| chr17 | 3542884 | 3542884 | SNV | C | C:T | 1 | | |
| chr17 | 3542895 | 3542895 | SNV | G | A:G | 1 | | |
| chr17 | 3542916 | 3542916 | SNV | C | C:T | 3 | | |
| chr17 | 3542991 | 3542991 | deletion | A | -:- | 51 | -:A | 149 |
| chr17 | 3543204 | 3543204 | SNV | C | C:T | 2 | | |

**Supplementary Table 1. Variants of *CTNS* in the 1KG database**

| Chromosome | Start Position | End Position | Variant Type | Reference Genotype \| Proteotype | Variant 1 Genotype \| Proteotype | Count | Variant 2 Genotype \| Proteotype | Count |
|---|---|---|---|---|---|---|---|---|
| chr17 | 3543271 | 3543271 | SNV | A | A:G | 442 | G:G | 170 |
| chr17 | 3543710 | 3543710 | SNV | G | A:G | 2 | | |
| chr17 | 3543855 | 3543855 | SNV | C | C:G | 1 | | |
| chr17 | 3543930 | 3543930 | SNV | G | C:G | 6 | | |
| chr17 | 3544017 | 3544017 | SNV | C | C:T | 358 | T:T | 123 |
| chr17 | 3544039 | 3544039 | deletion | T | -:- | 4 | -:T | 66 |
| chr17 | 3544054 | 3544054 | SNV | C | C:T | 31 | T:T | 2 |
| chr17 | 3544191 | 3544191 | SNV | C | A:A | 1 | A:C | 33 |
| chr17 | 3544209 | 3544209 | SNV | A | A:T | 3 | | |
| chr17 | 3544237 | 3544237 | SNV | T | A:T | 5 | | |
| chr17 | 3544295 | 3544295 | SNV | A | A:G | 442 | G:G | 169 |
| chr17 | 3544313 | 3544313 | SNV | G | A:A | 49 | A:G | 130 |
| chr17 | 3544405 | 3544405 | SNV | G | G:T | 13 | | |
| chr17 | 3544670 | 3544678 | deletion | ATATATATA | -:- | 161 | -:ATATATATA | 453 |
| chr17 | 3544679 | 3544679 | insertion | - | -:ATATATA | 267 | ATATATA:ATATATA | 56 |
| chr17 | 3544679 | 3544683 | deletion | TTTTT | -:- | 15 | -:TTTTT | 186 |
| chr17 | 3544729 | 3544729 | SNV | A | A:G | 443 | G:G | 172 |
| chr17 | 3544749 | 3544749 | SNV | G | A:G | 5 | | |
| chr17 | 3544779 | 3544779 | SNV | G | A:A | 171 | A:G | 441 |
| chr17 | 3544835 | 3544835 | SNV | G | A:A | 50 | A:G | 131 |
| chr17 | 3545030 | 3545030 | SNV | C | C:G | 5 | | |
| chr17 | 3545112 | 3545112 | SNV | G | A:A | 218 | A:G | 455 |
| chr17 | 3545127 | 3545127 | SNV | G | A:A | 171 | A:G | 440 |
| chr17 | 3545233 | 3545233 | SNV | G | A:A | 1 | A:G | 11 |
| chr17 | 3545302 | 3545302 | SNV | T | C:C | 218 | C:T | 455 |
| chr17 | 3545315 | 3545315 | SNV | G | A:A | 205 | A:G | 460 |
| chr17 | 3545323 | 3545323 | SNV | A | A:G | 458 | G:G | 215 |
| chr17 | 3545385 | 3545386 | deletion | GT | -:- | 9 | -:GT | 90 |
| chr17 | 3545416 | 3545416 | SNV | T | C:C | 1061 | C:T | 31 |
| chr17 | 3545515 | 3545515 | SNV | C | C:G | 1 | | |
| chr17 | 3545529 | 3545529 | SNV | A | A:G | 458 | G:G | 216 |
| chr17 | 3545559 | 3545559 | SNV | T | C:C | 219 | C:T | 454 |
| chr17 | 3545810 | 3545810 | SNV | C | C:T | 343 | T:T | 121 |
| chr17 | 3545811 | 3545811 | SNV | G | A:G | 25 | | |
| chr17 | 3545851 | 3545851 | SNV | A | A:G | 403 | G:G | 540 |
| chr17 | 3545873 | 3545873 | SNV | T | C:T | 16 | | |
| chr17 | 3546030 | 3546030 | SNV | C | C:T | 1 | | |
| chr17 | 3546093 | 3546093 | SNV | G | A:G | 2 | | |
| chr17 | 3546103 | 3546103 | SNV | A | A:G | 1 | | |
| chr17 | 3546130 | 3546130 | SNV | C | A:C | 1 | | |
| chr17 | 3546153 | 3546153 | SNV | T | C:C | 1 | C:T | 10 |
| chr17 | 3546160 | 3546160 | SNV | G | A:A | 123 | A:G | 355 |
| chr17 | 3546195 | 3546195 | SNV | C | C:G | 23 | G:G | 3 |
| chr17 | 3546231 | 3546231 | SNV | T | C:C | 1033 | C:T | 57 |
| chr17 | 3546347 | 3546347 | SNV | G | A:A | 216 | A:G | 456 |
| chr17 | 3546410 | 3546410 | SNV | T | C:C | 1 | C:T | 58 |
| chr17 | 3546424 | 3546424 | SNV | T | C:T | 6 | | |
| chr17 | 3546476 | 3546476 | SNV | C | C:T | 82 | T:T | 2 |

**Supplementary Table 1. Variants of *CTNS* in the 1KG database**

| Chromosome | Start Position | End Position | Variant Type | Reference Genotype \| Proteotype | Variant 1 Genotype \| Proteotype | Count | Variant 2 Genotype \| Proteotype | Count |
|---|---|---|---|---|---|---|---|---|
| chr17 | 3546557 | 3546557 | SNV | G | A:G | 1 | | |
| chr17 | 3546639 | 3546639 | SNV | T | C:C | 171 | C:T | 440 |
| chr17 | 3546665 | 3546665 | SNV | G | A:G | 28 | | |
| chr17 | 3546667 | 3546667 | SNV | T | C:T | 1 | | |
| chr17 | 3546887 | 3546890 | deletion | CTCA | -:- | 144 | -:CTCA | 379 |
| chr17 | 3546921 | 3546921 | SNV | C | C:T | 1 | | |
| chr17 | 3547105 | 3547105 | SNV | A | A:C | 467 | C:C | 229 |
| chr17 | 3547186 | 3547186 | SNV | T | C:C | 216 | C:T | 466 |
| chr17 | 3547187 | 3547187 | SNV | G | A:G | 1 | | |
| chr17 | 3547318 | 3547318 | SNV | T | G:T | 3 | | |
| chr17 | 3547451 | 3547451 | SNV | T | C:T | 1 | | |
| chr17 | 3547580 | 3547580 | SNV | G | A:G | 4 | | |
| chr17 | 3547583 | 3547583 | SNV | C | C:T | 7 | | |
| chr17 | 3547781 | 3547781 | insertion | - | -:T | 132 | T:T | 27 |
| chr17 | 3547832 | 3547832 | SNV | C | C:T | 1 | | |
| chr17 | 3547844 | 3547844 | SNV | C | C:T | 7 | | |
| chr17 | 3548199 | 3548199 | SNV | A | A:G | 130 | G:G | 49 |
| chr17 | 3548333 | 3548333 | SNV | T | C:T | 3 | | |
| chr17 | 3548447 | 3548447 | SNV | A | A:G | 468 | G:G | 229 |
| chr17 | 3548462 | 3548462 | SNV | C | A:C | 1 | | |
| chr17 | 3548617 | 3548617 | SNV | G | A:G | 1 | | |
| chr17 | 3548628 | 3548628 | SNV | T | G:T | 1 | | |
| chr17 | 3548647 | 3548647 | SNV | C | C:T | 467 | T:T | 230 |
| chr17 | 3548689 | 3548689 | SNV | G | A:A | 230 | A:G | 467 |
| chr17 | 3548720 | 3548720 | SNV | G | A:G | 2 | | |
| chr17 | 3548784 | 3548784 | SNV | C | C:T | 376 | T:T | 144 |
| chr17 | 3548806 | 3548806 | SNV | C | C:T | 1 | | |
| chr17 | 3548829 | 3548829 | SNV | G | A:A | 124 | A:G | 344 |
| chr17 | 3548981 | 3548981 | SNV | T | C:T | 18 | | |
| chr17 | 3549227 | 3549227 | SNV | G | C:C | 2 | C:G | 20 |
| chr17 | 3549239 | 3549239 | SNV | T | A:T | 14 | | |
| chr17 | 3549260 | 3549260 | deletion | A | -:- | 5 | -:A | 115 |
| chr17 | 3549267 | 3549267 | deletion | C | -:- | 55 | -:C | 257 |
| chr17 | 3549268 | 3549268 | SNV | C | A:A | 173 | A:C | 445 |
| chr17 | 3549270 | 3549270 | deletion | G | -:- | 183 | -:G | 453 |
| chr17 | 3549300 | 3549300 | SNV | G | A:A | 125 | A:G | 358 |
| chr17 | 3549405 | 3549405 | SNV | G | A:G | 1 | | |
| chr17 | 3549470 | 3549470 | SNV | T | C:C | 417 | C:T | 454 |
| chr17 | 3549543 | 3549543 | SNV | T | A:T | 3 | | |
| chr17 | 3549589 | 3549589 | insertion | - | -:C | 87 | C:C | 4 |
| chr17 | 3549615 | 3549615 | SNV | C | C:G | 35 | G:G | 2 |
| chr17 | 3549797 | 3549797 | SNV | G | A:G | 1 | | |
| chr17 | 3549826 | 3549826 | SNV | C | C:G | 31 | G:G | 1 |
| chr17 | 3549871 | 3549871 | SNV | A | A:G | 81 | G:G | 1 |
| chr17 | 3550082 | 3550082 | insertion | - | -:G | 122 | G:G | 870 |
| chr17 | 3550133 | 3550133 | SNV | G | A:G | 1 | | |
| chr17 | 3550163 | 3550163 | SNV | T | A:A | 2 | A:T | 12 |
| chr17 | 3550252 | 3550252 | SNV | C | C:G | 1 | | |

**Supplementary Table 1. Variants of *CTNS* in the 1KG database**

| Chromosome | Start Position | End Position | Variant Type | Reference Genotype \| Proteotype | Variant 1 Genotype \| Proteotype | Count | Variant 2 Genotype \| Proteotype | Count |
|---|---|---|---|---|---|---|---|---|
| chr17 | 3550284 | 3550284 | SNV | C | C:G | 101 | G:G | 5 |
| chr17 | 3550396 | 3550396 | SNV | C | C:T | 101 | T:T | 5 |
| chr17 | 3550408 | 3550408 | SNV | A | A:G | 134 | G:G | 899 |
| chr17 | 3550409 | 3550409 | SNV | C | C:T | 12 | | |
| chr17 | 3550514 | 3550514 | SNV | C | C:T | 73 | T:T | 2 |
| chr17 | 3550520 | 3550520 | SNV | G | A:G | 1 | | |
| chr17 | 3550784 | 3550784 | SNV | C\|N | C:T \| N:N | 8 | T:T \| N:N | 1 |
| chr17 | 3550792 | 3550792 | SNV | C\|S | C:T \| S:L | 2 | | |
| chr17 | 3550799 | 3550799 | SNV | C\|N | C:T \| N:N | 3 | | |
| chr17 | 3550800 | 3550800 | SNV | G\|V | A:G \| I:V | 35 | | |
| chr17 | 3550842 | 3550842 | SNV | C | C:T | 1 | | |
| chr17 | 3550850 | 3550850 | SNV | C | C:T | 1 | | |
| chr17 | 3551123 | 3551123 | SNV | C | C:T | 5 | | |
| chr17 | 3551178 | 3551178 | SNV | A | A:G | 154 | G:G | 835 |
| chr17 | 3551199 | 3551199 | deletion | T | -:- | 2 | -:T | 44 |
| chr17 | 3551310 | 3551310 | SNV | C | C:T | 284 | T:T | 41 |
| chr17 | 3551327 | 3551327 | SNV | T | C:C | 863 | C:T | 166 |
| chr17 | 3551345 | 3551345 | SNV | T | G:G | 124 | G:T | 358 |
| chr17 | 3551586 | 3551586 | SNV | C | C:T | 3 | | |
| chr17 | 3551616 | 3551616 | SNV | T | C:C | 1092 | | |
| chr17 | 3551799 | 3551799 | SNV | C | C:T | 5 | | |
| chr17 | 3551981 | 3551981 | SNV | C | C:T | 2 | | |
| chr17 | 3551994 | 3551994 | SNV | T | G:T | 1 | | |
| chr17 | 3551998 | 3551998 | SNV | C | C:T | 2 | | |
| chr17 | 3552038 | 3552038 | SNV | G | A:A | 144 | A:G | 375 |
| chr17 | 3552261 | 3552261 | SNV | G | A:G | 1 | | |
| chr17 | 3552372 | 3552372 | SNV | G | C:C | 124 | C:G | 357 |
| chr17 | 3552388 | 3552388 | SNV | G | A:G | 2 | | |
| chr17 | 3552389 | 3552389 | SNV | C | A:C | 2 | | |
| chr17 | 3552466 | 3552466 | SNV | G | A:G | 3 | | |
| chr17 | 3552532 | 3552532 | SNV | A | A:G | 52 | G:G | 8 |
| chr17 | 3552678 | 3552678 | SNV | G | A:G | 3 | | |
| chr17 | 3552727 | 3552727 | SNV | C | C:T | 281 | T:T | 40 |
| chr17 | 3552815 | 3552815 | SNV | A | A:T | 1 | | |
| chr17 | 3552890 | 3552890 | SNV | A | A:G | 16 | | |
| chr17 | 3553012 | 3553012 | SNV | C | A:C | 16 | | |
| chr17 | 3553056 | 3553056 | insertion | - | -:C | 427 | C:C | 248 |
| chr17 | 3553058 | 3553058 | insertion | - | -:T | 407 | T:T | 278 |
| chr17 | 3553148 | 3553148 | SNV | C | C:T | 1 | | |
| chr17 | 3553270 | 3553270 | SNV | A | A:G | 16 | | |
| chr17 | 3553292 | 3553292 | SNV | A | A:C | 42 | C:C | 1 |
| chr17 | 3553358 | 3553358 | SNV | A | A:G | 1 | | |
| chr17 | 3553426 | 3553426 | SNV | A | A:T | 1 | | |
| chr17 | 3553430 | 3553430 | SNV | A | A:G | 16 | | |
| chr17 | 3553451 | 3553451 | SNV | A | A:G | 3 | | |
| chr17 | 3553474 | 3553474 | SNV | G | A:G | 16 | | |
| chr17 | 3553477 | 3553477 | SNV | G | A:G | 1 | | |
| chr17 | 3553489 | 3553489 | SNV | G | A:G | 1 | | |

**Supplementary Table 1. Variants of *CTNS* in the 1KG database**

| Chromosome | Start Position | End Position | Variant Type | Reference Genotype \| Proteotype | Variant 1 Genotype \| Proteotype | Count | Variant 2 Genotype \| Proteotype | Count |
|---|---|---|---|---|---|---|---|---|
| chr17 | 3553526 | 3553526 | SNV | A | A:G | 166 | G:G | 863 |
| chr17 | 3553582 | 3553582 | SNV | G | A:G | 19 | | |
| chr17 | 3553655 | 3553655 | SNV | A | A:G | 85 | G:G | 5 |
| chr17 | 3553665 | 3553665 | SNV | A | A:G | 37 | G:G | 1 |
| chr17 | 3553743 | 3553743 | SNV | T | C:C | 2 | C:T | 10 |
| chr17 | 3553766 | 3553766 | SNV | A | A:C | 12 | | |
| chr17 | 3554009 | 3554009 | SNV | G | A:A | 1 | A:G | 37 |
| chr17 | 3554059 | 3554059 | SNV | G | A:A | 375 | A:G | 462 |
| chr17 | 3554136 | 3554136 | SNV | G | A:G | 2 | | |
| chr17 | 3554182 | 3554182 | SNV | T | C:T | 16 | | |
| chr17 | 3554295 | 3554295 | SNV | C | C:T | 15 | | |
| chr17 | 3554552 | 3554552 | SNV | C | C:G | 37 | G:G | 1 |
| chr17 | 3554631 | 3554631 | SNV | C | C:T | 14 | T:T | 1 |
| chr17 | 3554669 | 3554669 | SNV | G | A:A | 831 | A:G | 146 |
| chr17 | 3554721 | 3554721 | SNV | A | A:G | 6 | | |
| chr17 | 3554812 | 3554812 | SNV | C | C:T | 10 | T:T | 1 |
| chr17 | 3554817 | 3554817 | SNV | G | A:A | 1 | A:G | 44 |
| chr17 | 3554941 | 3554941 | SNV | A | A:G | 2 | | |
| chr17 | 3554993 | 3554993 | SNV | C | C:T | 8 | | |
| chr17 | 3554994 | 3554994 | SNV | C | C:G | 160 | G:G | 863 |
| chr17 | 3554997 | 3554997 | deletion | G | -:- | 2 | -:G | 76 |
| chr17 | 3555153 | 3555153 | SNV | A | A:G | 2 | | |
| chr17 | 3555277 | 3555277 | deletion | G | -:- | 843 | -:G | 182 |
| chr17 | 3555288 | 3555288 | SNV | A | A:T | 16 | | |
| chr17 | 3555289 | 3555289 | SNV | A | A:C | 16 | | |
| chr17 | 3555390 | 3555390 | SNV | A | A:C | 16 | | |
| chr17 | 3555424 | 3555424 | SNV | G | A:A | 40 | A:G | 281 |
| chr17 | 3555439 | 3555439 | SNV | G | A:G | 13 | | |
| chr17 | 3555492 | 3555492 | SNV | A | A:C | 6 | | |
| chr17 | 3555624 | 3555624 | SNV | G | C:G | 4 | | |
| chr17 | 3555652 | 3555652 | SNV | G | A:A | 2 | A:G | 80 |
| chr17 | 3555747 | 3555747 | SNV | G | A:G | 3 | | |
| chr17 | 3555754 | 3555754 | SNV | C | C:T | 1 | | |
| chr17 | 3555817 | 3555817 | SNV | C | C:T | 172 | T:T | 843 |
| chr17 | 3555867 | 3555867 | SNV | C | C:T | 4 | | |
| chr17 | 3555894 | 3555894 | SNV | A | A:T | 3 | | |
| chr17 | 3555915 | 3555915 | SNV | C | C:T | 3 | | |
| chr17 | 3555986 | 3555986 | SNV | T | C:C | 863 | C:T | 166 |
| chr17 | 3556120 | 3556120 | SNV | T | C:T | 16 | | |
| chr17 | 3556140 | 3556140 | SNV | T | C:T | 16 | | |
| chr17 | 3556150 | 3556150 | insertion | - | -:C | 148 | C:C | 9 |
| chr17 | 3556292 | 3556292 | SNV | G | A:G | 3 | | |
| chr17 | 3556324 | 3556324 | SNV | C | C:T | 3 | | |
| chr17 | 3556432 | 3556432 | SNV | A | G:G | 1092 | | |
| chr17 | 3556456 | 3556456 | SNV | A | A:G | 27 | G:G | 2 |
| chr17 | 3556463 | 3556463 | SNV | T | C:T | 16 | | |
| chr17 | 3556471 | 3556471 | SNV | C | C:T | 14 | | |
| chr17 | 3556494 | 3556494 | SNV | T | C:T | 1 | | |

**Supplementary Table 1. Variants of *CTNS* in the 1KG database**

| Chromosome | Start Position | End Position | Variant Type | Reference Genotype \| Proteotype | Variant 1 Genotype \| Proteotype | Count | Variant 2 Genotype \| Proteotype | Count |
|---|---|---|---|---|---|---|---|---|
| chr17 | 3556498 | 3556498 | SNV | A | A:G | 1 | | |
| chr17 | 3556501 | 3556501 | SNV | A | A:T | 1 | | |
| chr17 | 3556504 | 3556504 | SNV | G | A:G | 1 | | |
| chr17 | 3556563 | 3556563 | SNV | T | C:C | 861 | C:T | 170 |
| chr17 | 3556594 | 3556594 | SNV | C | C:T | 313 | T:T | 108 |
| chr17 | 3556600 | 3556600 | SNV | A | A:G | 16 | | |
| chr17 | 3556623 | 3556623 | SNV | C | C:G | 12 | | |
| chr17 | 3556782 | 3556782 | SNV | A | A:G | 3 | | |
| chr17 | 3556832 | 3556832 | SNV | G | A:G | 1 | | |
| chr17 | 3556897 | 3556897 | SNV | C | C:T | 358 | T:T | 142 |
| chr17 | 3556920 | 3556920 | SNV | A | A:G | 16 | | |
| chr17 | 3556983 | 3556983 | SNV | A | A:G | 2 | | |
| chr17 | 3557042 | 3557042 | SNV | C | C:T | 1 | | |
| chr17 | 3557055 | 3557055 | SNV | A | A:T | 7 | | |
| chr17 | 3557076 | 3557076 | SNV | G | A:G | 1 | | |
| chr17 | 3557131 | 3557131 | SNV | T | C:T | 19 | | |
| chr17 | 3557175 | 3557175 | insertion | - | -:A | 95 | A:A | 978 |
| chr17 | 3557181 | 3557181 | deletion | T | -:- | 861 | -:T | 168 |
| chr17 | 3557219 | 3557219 | SNV | T | C:T | 7 | | |
| chr17 | 3557381 | 3557381 | SNV | A | A:T | 1 | | |
| chr17 | 3557382 | 3557382 | SNV | A | A:T | 164 | T:T | 867 |
| chr17 | 3557495 | 3557495 | SNV | A | A:G | 387 | G:G | 145 |
| chr17 | 3557499 | 3557499 | SNV | C | C:T | 345 | T:T | 123 |
| chr17 | 3557537 | 3557537 | SNV | C | C:T | 158 | T:T | 859 |
| chr17 | 3557594 | 3557594 | SNV | A | A:G | 158 | G:G | 859 |
| chr17 | 3557601 | 3557601 | SNV | C | C:T | 13 | T:T | 1 |
| chr17 | 3557647 | 3557647 | SNV | G | G:T | 1 | | |
| chr17 | 3557837 | 3557837 | SNV | G | A:G | 4 | | |
| chr17 | 3557846 | 3557846 | SNV | T | G:G | 1 | G:T | 34 |
| chr17 | 3557890 | 3557890 | SNV | G | A:G | 1 | | |
| chr17 | 3558024 | 3558024 | SNV | C | C:T | 1 | | |
| chr17 | 3558098 | 3558098 | SNV | G | A:G | 16 | | |
| chr17 | 3558185 | 3558185 | SNV | C | C:T | 11 | T:T | 1 |
| chr17 | 3558212 | 3558212 | SNV | C | A:C | 3 | | |
| chr17 | 3558266 | 3558266 | SNV | G | A:G | 2 | | |
| chr17 | 3558417 | 3558417 | SNV | C | C:G | 166 | G:G | |
| chr17 | 3558426 | 3558426 | SNV | C | A:C | 1 | | |
| chr17 | 3558443 | 3558443 | SNV | C | C:T | 4 | | |
| chr17 | 3558467 | 3558467 | SNV | T | C:T | 2 | | |
| chr17 | 3558486 | 3558486 | SNV | C | C:T | 1 | | |
| chr17 | 3558549 | 3558549 | SNV | G\|A | A:G \| T:A | 1 | | |
| chr17 | 3558553 | 3558553 | SNV | T\|I | G:T \| S:I | 1 | | |
| chr17 | 3558688 | 3558688 | SNV | T | C:T | 1 | | |
| chr17 | 3558695 | 3558695 | SNV | A | A:G | 386 | G:G | 145 |
| chr17 | 3558698 | 3558698 | SNV | G | A:A | 378 | A:G | 472 |
| chr17 | 3558700 | 3558700 | SNV | G | A:A | 46 | A:G | 301 |
| chr17 | 3558760 | 3558760 | SNV | A | A:G | 3 | | |
| chr17 | 3558963 | 3558963 | SNV | C | A:A | 4 | A:C | 85 |

**Supplementary Table 1. Variants of *CTNS* in the 1KG database**

| Chromosome | Start Position | End Position | Variant Type | Reference Genotype \| Proteotype | Variant 1 Genotype \| Proteotype | Count | Variant 2 Genotype \| Proteotype | Count |
|---|---|---|---|---|---|---|---|---|
| chr17 | 3559273 | 3559273 | SNV | G | G:T | 5 | T:T | 1 |
| chr17 | 3559303 | 3559303 | SNV | T | G:G | 2 | G:T | 57 |
| chr17 | 3559430 | 3559430 | SNV | G | A:A | 2 | A:G | 89 |
| chr17 | 3559451 | 3559451 | SNV | T | C:C | 12 | C:T | 199 |
| chr17 | 3559481 | 3559481 | SNV | A | A:C | 235 | C:C | 18 |
| chr17 | 3559502 | 3559502 | SNV | T | C:C | 100 | C:T | 353 |
| chr17 | 3559506 | 3559506 | SNV | T | C:C | 101 | C:T | 350 |
| chr17 | 3559511 | 3559511 | SNV | A | A:C | 403 | C:C | 88 |
| chr17 | 3559517 | 3559517 | SNV | C | A:A | 137 | A:C | 347 |
| chr17 | 3559651 | 3559651 | SNV | C | C:T | 235 | T:T | 42 |
| chr17 | 3559676 | 3559677 | deletion | CT | -:- | 20 | -:CT | 169 |
| chr17 | 3559687 | 3559687 | SNV | C | C:T | 246 | T:T | 22 |
| chr17 | 3559722 | 3559722 | SNV | T | C:T | 5 | | |
| chr17 | 3559742 | 3559742 | SNV | T | C:T | 1 | | |
| chr17 | 3559774 | 3559774 | SNV | C | A:C | 7 | | |
| chr17 | 3559781 | 3559781 | SNV | T\|S | C:C \| S:S | | C:T \| S:S | 89 |
| chr17 | 3559792 | 3559792 | SNV | T\|L | C:T \| P:L | 1 | | |
| chr17 | 3559806 | 3559806 | SNV | G\|V | A:G \| M:V | 1 | | |
| chr17 | 3559823 | 3559823 | SNV | G\|T | A:A \| T:T | 39 | A:G \| T:T | 281 |
| chr17 | 3559884 | 3559884 | SNV | C | C:T | 1 | | |
| chr17 | 3559901 | 3559901 | SNV | C | A:C | 1 | | |
| chr17 | 3560009 | 3560009 | SNV | G\|V | A:G \| M:V | 1 | | |
| chr17 | 3560020 | 3560020 | SNV | C\|N | C:T \| N:N | 2 | | |
| chr17 | 3560110 | 3560110 | SNV | C | A:C | 2 | | |
| chr17 | 3560363 | 3560363 | SNV | G | A:A | 47 | A:G | 306 |
| chr17 | 3560371 | 3560377 | deletion | AGGGAGG | -:- | 11 | -:AGGGAGG | 115 |
| chr17 | 3560450 | 3560450 | SNV | G | A:G | 1 | | |
| chr17 | 3560541 | 3560542 | deletion | GA | -:- | 5 | -:GA | 30 |
| chr17 | 3560561 | 3560561 | SNV | G | G:T | 3 | | |
| chr17 | 3560598 | 3560598 | SNV | G | A:A | 50 | A:G | 129 |
| chr17 | 3560683 | 3560683 | SNV | G | A:A | 13 | A:G | 67 |
| chr17 | 3560752 | 3560752 | SNV | T | C:C | 1030 | C:T | 61 |
| chr17 | 3560987 | 3560987 | SNV | G | A:G | 2 | | |
| chr17 | 3561200 | 3561200 | SNV | C | C:T | 1 | | |
| chr17 | 3561201 | 3561201 | SNV | G | A:A | 3 | A:G | 54 |
| chr17 | 3561233 | 3561233 | SNV | C | A:A | 3 | A:C | 54 |
| chr17 | 3561249 | 3561249 | SNV | C | C:T | 2 | | |
| chr17 | 3561302 | 3561302 | SNV | G\|G | A:G \| S:G | 2 | | |
| chr17 | 3561312 | 3561312 | SNV | G\|R | A:G \| H:R | 2 | | |
| chr17 | 3561396 | 3561396 | SNV | C\|T | C:T \| T:I | 210 | T:T \| I:I | 812 |
| chr17 | 3561451 | 3561451 | SNV | G\|L | A:G \| L:L | 1 | | |
| chr17 | 3561671 | 3561671 | SNV | G | A:G | 3 | | |
| chr17 | 3561697 | 3561697 | SNV | G | A:A | 1 | A:G | 59 |
| chr17 | 3561711 | 3561711 | SNV | C | C:G | 404 | G:G | 434 |
| chr17 | 3561712 | 3561712 | SNV | A | A:G | 402 | G:G | 436 |
| chr17 | 3561725 | 3561725 | SNV | G | A:G | 3 | | |
| chr17 | 3561728 | 3561728 | SNV | C | C:G | 176 | G:G | 61 |
| chr17 | 3561744 | 3561744 | SNV | C | C:T | 16 | | |

**Supplementary Table 1. Variants of *CTNS* in the 1KG database**

| Chromosome | Start Position | End Position | Variant Type | Reference Genotype \| Proteotype | Variant 1 Genotype \| Proteotype | Count | Variant 2 Genotype \| Proteotype | Count |
|---|---|---|---|---|---|---|---|---|
| chr17 | 3561750 | 3561750 | SNV | T | C:C | 56 | C:T | 177 |
| chr17 | 3561765 | 3561765 | SNV | G | A:G | 3 | | |
| chr17 | 3561817 | 3561817 | SNV | G | A:G | 21 | | |
| chr17 | 3561842 | 3561842 | SNV | C | C:T | 448 | T:T | 145 |
| chr17 | 3561912 | 3561912 | SNV | A | A:T | 6 | | |
| chr17 | 3561956 | 3561956 | SNV | C | C:T | 348 | T:T | 52 |
| chr17 | 3561961 | 3561961 | SNV | G | A:A | 1 | A:G | 5 |
| chr17 | 3562040 | 3562040 | SNV | C | C:G | 376 | G:G | 140 |
| chr17 | 3562182 | 3562182 | SNV | G | A:A | 975 | A:G | 104 |
| chr17 | 3562196 | 3562196 | SNV | G | A:A | 52 | A:G | 335 |
| chr17 | 3562200 | 3562200 | SNV | T | C:C | 50 | C:T | 339 |
| chr17 | 3562202 | 3562202 | SNV | T | C:C | 50 | C:T | 334 |
| chr17 | 3562405 | 3562405 | SNV | G | A:G | 11 | | |
| chr17 | 3562485 | 3562485 | insertion | - | -:C | 481 | C:C | 260 |
| chr17 | 3562486 | 3562486 | insertion | - | -:G | 412 | G:G | 425 |
| chr17 | 3562488 | 3562488 | insertion | - | -:C | 496 | C:C | 228 |
| chr17 | 3562573 | 3562573 | SNV | C | C:T | 3 | | |
| chr17 | 3562617 | 3562617 | SNV | C | C:T | 2 | | |
| chr17 | 3562624 | 3562624 | SNV | G | G:T | 57 | T:T | 3 |
| chr17 | 3562818 | 3562819 | deletion | AT | -:- | 3 | -:AT | 73 |
| chr17 | 3562916 | 3562916 | SNV | G | A:G | 7 | | |
| chr17 | 3563009 | 3563009 | SNV | C | C:T | 1 | | |
| chr17 | 3563091 | 3563091 | SNV | G | A:G | 2 | | |
| chr17 | 3563124 | 3563124 | SNV | C | C:T | 10 | | |
| chr17 | 3563125 | 3563125 | SNV | G | A:G | 2 | | |
| chr17 | 3563196 | 3563196 | SNV | T\|I | C:T \| I:I | 1 | | |
| chr17 | 3563220 | 3563220 | SNV | C\|T | C:T \| T:T | 1 | | |
| chr17 | 3563284 | 3563284 | SNV | G | A:A | 1 | A:G | 49 |
| chr17 | 3563314 | 3563314 | SNV | C | C:T | 131 | T:T | 48 |
| chr17 | 3563339 | 3563339 | SNV | C | C:T | 373 | T:T | 138 |
| chr17 | 3563416 | 3563416 | SNV | G | A:G | 3 | | |
| chr17 | 3563550 | 3563550 | SNV | G\|G | A:G \| R:G | 1 | | |
| chr17 | 3563588 | 3563588 | SNV | C\|I | C:T \| I:I | 1 | | |
| chr17 | 3563743 | 3563743 | SNV | G | A:G | 1 | | |
| chr17 | 3563923 | 3563923 | SNV | G\|A | G:T \| A:A | 1 | | |
| chr17 | 3563925 | 3563925 | SNV | G\|R | A:G \| H:R | 2 | | |
| chr17 | 3563963 | 3563963 | SNV | C\|P | C:G \| P:A | 329 | G:G \| A:A | 104 |
| chr17 | 3563976 | 3563976 | SNV | C\|P | C:T \| P:L | 2 | | |
| chr17 | 3563992 | 3563992 | SNV | A\|Q | A:G \| Q:Q | 3 | | |
| chr17 | 3564037 | 3564037 | SNV | C | C:T | 1 | | |
| chr17 | 3564068 | 3564068 | SNV | G | A:A | 52 | A:G | 347 |
| chr17 | 3564073 | 3564073 | SNV | A | A:G | 348 | G:G | 52 |
| chr17 | 3564084 | 3564084 | SNV | T | C:C | 52 | C:T | 347 |
| chr17 | 3564132 | 3564132 | SNV | G | A:G | 1 | | |
| chr17 | 3564190 | 3564190 | SNV | G | A:A | 12 | A:G | 83 |
| chr17 | 3564218 | 3564218 | SNV | T | C:C | 52 | C:T | 347 |
| chr17 | 3564235 | 3564235 | SNV | A | A:G | 347 | G:G | 52 |
| chr17 | 3564290 | 3564290 | SNV | T | C:C | 52 | C:T | 346 |

**Supplementary Table 1. Variants of *CTNS* in the 1KG database**

| Chromosome | Start Position | End Position | Variant Type | Reference Genotype \| Proteotype | Variant 1 Genotype \| Proteotype | Count | Variant 2 Genotype \| Proteotype | Count |
|---|---|---|---|---|---|---|---|---|
| chr17 | 3564294 | 3564294 | SNV | T | C:C | 52 | C:T | 346 |
| chr17 | 3564351 | 3564351 | SNV | G | A:G | 1 | | |
| chr17 | 3564378 | 3564378 | SNV | C | C:T | 346 | T:T | 51 |
| chr17 | 3564385 | 3564385 | SNV | C | C:T | 1 | | |
| chr17 | 3564400 | 3564401 | deletion | TT | -:- | 83 | -:TT | 268 |
| chr17 | 3564400 | 3564400 | SNV | T | A:A | 125 | A:T | 359 |
| chr17 | 3564401 | 3564401 | SNV | T | G:G | 1086 | G:T | 6 |
| chr17 | 3564498 | 3564498 | SNV | A | A:C | 6 | | |
| chr17 | 3564510 | 3564510 | SNV | C | C:T | 8 | | |
| chr17 | 3564514 | 3564514 | SNV | G | A:A | 3 | A:G | 29 |
| chr17 | 3564605 | 3564605 | SNV | C | C:T | 4 | | |
| chr17 | 3564660 | 3564660 | SNV | C | C:T | 1 | | |
| chr17 | 3564687 | 3564687 | SNV | C | C:T | 288 | T:T | 43 |
| chr17 | 3564688 | 3564688 | SNV | A | A:G | 288 | G:G | 42 |
| chr17 | 3564693 | 3564693 | insertion | - | -:GTC | 290 | GTC:GTC | 45 |
| chr17 | 3564708 | 3564708 | insertion | - | -:G | 132 | G:G | 7 |
| chr17 | 3564711 | 3564711 | insertion | - | -:G | 211 | G:G | 24 |
| chr17 | 3564716 | 3564716 | insertion | - | -:G | 224 | G:G | 25 |
| chr17 | 3564753 | 3564753 | SNV | T | C:T | 1 | | |
| chr17 | 3564779 | 3564779 | SNV | C | C:T | 329 | T:T | 52 |
| chr17 | 3564780 | 3564780 | SNV | G | A:A | 1 | A:G | 32 |
| chr17 | 3564791 | 3564791 | SNV | G | A:G | 11 | | |
| chr17 | 3564815 | 3564815 | SNV | A | A:G | 327 | G:G | |
| chr17 | 3564823 | 3564823 | SNV | G | A:G | 42 | | |
| chr17 | 3564828 | 3564828 | SNV | C | C:T | 42 | T:T | 1 |
| chr17 | 3564837 | 3564838 | deletion | CT | -:- | 52 | -:CT | 329 |
| chr17 | 3564869 | 3564869 | SNV | T | C:C | 2 | C:T | 12 |
| chr17 | 3564945 | 3564945 | SNV | C | C:T | 1 | | |
| chr17 | 3564946 | 3564946 | SNV | G | A:G | 3 | | |
| chr17 | 3564951 | 3564951 | SNV | G | A:G | 3 | | |
| chr17 | 3565103 | 3565103 | SNV | A | A:G | 398 | G:G | 585 |
| chr17 | 3565144 | 3565144 | SNV | T | C:T | 478 | | |
| chr17 | 3565241 | 3565241 | SNV | C | C:G | 32 | G:G | 1 |
| chr17 | 3565258 | 3565258 | SNV | C | C:T | 56 | T:T | 1 |
| chr17 | 3565281 | 3565281 | SNV | C | C:G | 2 | | |
| chr17 | 3565296 | 3565296 | SNV | C | C:G | 56 | G:G | 1 |
| chr17 | 3565328 | 3565328 | SNV | A | A:G | 72 | G:G | 1 |
| chr17 | 3565331 | 3565331 | SNV | G | A:G | 1 | | |
| chr17 | 3565332 | 3565332 | SNV | C | C:T | 15 | | |
| chr17 | 3565408 | 3565408 | SNV | C | C:T | 1 | | |
| chr17 | 3565474 | 3565474 | SNV | G | A:G | 10 | | |
| chr17 | 3565529 | 3565529 | SNV | C | C:T | 54 | T:T | 1 |
| chr17 | 3565622 | 3565622 | SNV | T | C:T | 6 | | |
| chr17 | 3565675 | 3565675 | SNV | C | C:T | 10 | T:T | 1 |
| chr17 | 3565699 | 3565699 | SNV | G | A:G | 3 | | |
| chr17 | 3565706 | 3565706 | SNV | G | G:T | 5 | | |
| chr17 | 3565745 | 3565745 | SNV | C | C:T | 4 | | |
| chr17 | 3565778 | 3565778 | SNV | G | A:A | 1 | A:G | 54 |

**Supplementary Table 1. Variants of *CTNS* in the 1KG database**

| Chromosome | Start Position | End Position | Variant Type | Reference Genotype \| Proteotype | Variant 1 Genotype \| Proteotype | Count | Variant 2 Genotype \| Proteotype | Count |
|---|---|---|---|---|---|---|---|---|
| chr17 | 3565782 | 3565782 | SNV | G | G:T | 6 | | |
| chr17 | 3565790 | 3565790 | SNV | G | A:A | 1 | A:G | 24 |
| chr17 | 3565795 | 3565795 | SNV | G | A:G | 6 | | |
| chr17 | 3565872 | 3565873 | deletion | GG | -:- | 1 | -:GG | 23 |
| chr17 | 3565964 | 3565964 | SNV | C | C:T | 3 | | |
| chr17 | 3565969 | 3565969 | SNV | T | C:C | 554 | C:T | 370 |
| chr17 | 3566021 | 3566021 | SNV | T | A:T | 1 | | |
| chr17 | 3566045 | 3566045 | insertion | - | -:C | 334 | C:C | 690 |
| chr17 | 3566066 | 3566066 | SNV | T | G:T | 1 | | |
| chr17 | 3566209 | 3566209 | SNV | G | C:C | 40 | C:G | 271 |
| chr17 | 3566213 | 3566213 | SNV | G | A:A | 40 | A:G | 271 |
| chr17 | 3566232 | 3566232 | SNV | C | C:T | 460 | T:T | 164 |
